# Occupational burnout in the UK film and television industry: A cross-sectional survey of mental health, job quality, perceived stigma, and sexual harassment

**DOI:** 10.64898/2026.10.15.26365611

**Authors:** Mark Caldwell, Rebecca Chamberlain, Daniel Rippon

## Abstract

Recent industry surveys have reported widespread concern about mental health, working conditions, and harassment in the UK film and television industry, but no peer-reviewed empirical study has examined the determinants of occupational burnout in this workforce. The present study tested whether physical health, mental wellbeing, perceived public stigma, intrinsic job quality, and sexual harassment exposure predict burnout in industry professionals. A cross-sectional online survey was completed by 415 individuals across more than 100 job titles, with 286 also completing an optional harassment module. Mental wellbeing sat well below population norms, with 68% scoring in the range indicative of probable depressive symptoms and 82% in the high-exhaustion range on the Maslach Burnout Inventory General Survey. Mental wellbeing and intrinsic job quality were the strongest predictors of burnout (R² = .56), with meaningfulness accounting for most of the job-quality effect. Working hours independently predicted emotional exhaustion. Harassment exposure was associated with burnout in women but not men, approximately 30% mediated by mental wellbeing; sexist behaviour directly predicted exhaustion in women. Workers with a disability reported higher stigma. Burnout was strongly associated with intent to leave, and the findings collectively indicate an industry-wide occupational health concern requiring structural rather than individual-level intervention.

## Introduction

The United Kingdom film and television industry is among the world’s leading hubs for film and high-end television production [1], but recent industry surveys have raised mounting concern about the wellbeing of the workforce sustaining it. The Film and TV Charity’s Looking Glass surveys reported that the majority of respondents described their mental health as poor or below average and that bullying, harassment, and stigmatising attitudes about mental ill-health were widespread [2,3]. BECTU’s Eyes Half Shut survey documented routinely long working days, chronic fatigue, suicidal ideation, and a culture that normalises overwork [4]. Equity’s scoping review of the performing arts identified comparable patterns [5], and a Film and TV Charity retention study concluded that workers were leaving the industry in unsustainable numbers [6]. While invaluable for raising the visibility of an underexamined workforce, this grey literature is descriptive rather than analytic. Peer-reviewed research on the mental health of UK creative industry workers has focused primarily on the performing arts [7] and the cultural sector at large [8], with pandemic-era conditions as the analytic anchor. Occupational burnout in the UK film and television workforce, and the psychosocial predictors of it under ordinary industry working conditions, remains unexamined. The present study tests which psychosocial factors most strongly predict occupational burnout in this workforce.

Occupational burnout is a psychological syndrome of emotional exhaustion, cynicism about the value of one’s work, and reduced professional efficacy, developing in response to chronic interpersonal and organisational stressors [9,10]. Burnout predicts absenteeism, turnover, depression, cardiovascular disease, and increased suicide risk [11,12]. It has been studied extensively in healthcare, education, and human services, but rarely in creative occupations, where workers are typically self-employed, employment is project-based, and the organisational structures that buffer occupational stressors are often weakened or absent [13,14].

Two theoretical frameworks organise the predictors we examined. The Job Demands-Resources framework (JD-R) [15] proposes that burnout emerges when workplace demands exceed available resources, with demands including work intensity, long hours, and interpersonal stressors, and resources including autonomy, meaningfulness, and colleague interaction. The Conservation of Resources theory (COR) [16] complements this by proposing that workers who cannot recover between demand episodes become especially vulnerable. Both frameworks were developed largely in salaried samples with stable employment, making the predominantly freelance, project-based UK film and television workforce a distinctive test case. The predictors examined below map onto the JD-R demand-resource distinction, with intrinsic job quality operationalised through the European Intrinsic Job Quality Index [17].

Occupational burnout is not uniformly distributed across a workforce. A meta-analysis of 409 effect sizes across 183 studies [18] found small but consistent gender differences: women reported slightly higher emotional exhaustion than men (δ = 0.10), while men reported slightly higher cynicism or depersonalisation (δ = –0.19). Occupational gender typing did not moderate these differences meta-analytically, indicating the pattern is not merely an artefact of women concentrating in emotionally demanding sectors. Industry surveys of UK film and television have consistently reported that women in the workforce face sexual harassment and inequitable treatment [3,5], and the feminist critique of creative labour has identified structural mechanisms through which women’s careers in these industries are systematically constrained [19,20]. We therefore examined gender differences in each measured variable as secondary analyses and treated sexual harassment exposure as a formal predictor hypothesis.

We considered four features of industry work as primary predictors in the full sample, and a fifth, sexual harassment exposure, as a planned secondary analysis on the subsample completing an optional module. Each predictor maps onto recurring concerns in the industry surveys and was operationalised through an established psychometric instrument.

### Physical health

Long working hours, shift work, and time pressure compromise physical health and elevate the risk of cardiovascular, metabolic, and musculoskeletal conditions [21,22], and poor physical health is associated with elevated burnout through reduced recovery capacity and bidirectional links to mental wellbeing [23,24]. We expected lower physical health to be associated with higher burnout (Hypothesis 1).

### Mental wellbeing

Burnout overlaps conceptually and empirically with depression and general mental ill-health, though the constructs are not synonymous [25,26]. Workers with chronic low mood have fewer cognitive and emotional resources for job demands, accelerating the exhaustion and cynicism that characterise burnout [27,28]. Given the widespread mental ill-health reported in the industry [3], we expected lower mental wellbeing to be associated with higher burnout (Hypothesis 2).

### Perceived public stigma

Perceived public stigma about mental ill-health can operate as an additional chronic demand for workers, deterring help-seeking and imposing the ongoing stressor of concealment [29,30] and is linked to greater distress and reduced help-seeking in working populations [31,32]. We measured perceived public stigma, that is, expectations of negative beliefs and behaviour towards mentally ill people, using the STIG-9 [33], and expected higher stigma to be associated with higher burnout (Hypothesis 3).

### Intrinsic job quality

Burnout has been consistently linked to imbalances between job demands and resources [10,34]. The European Intrinsic Job Quality Index (EIJQI) [17] integrates four such subdimensions: autonomy, interaction with colleagues, intensity, and meaningfulness, each mapping onto recurring industry concerns about working hours, leadership support, and workplace bullying [3–5]. We expected lower intrinsic job quality to be associated with higher burnout (Hypothesis 4), and as a secondary aim examined which subdimension most strongly accounted for the relationship.

### Sexual harassment exposure

Concern about sexual harassment in entertainment industries came to global prominence with the #MeToo movement [35,36] and UK industry reports have documented sustained harassment concerns in film and television [3,19,37]. Across occupational sectors, harassment exposure is associated with elevated job stress, distress, and burnout, with higher exposure reported by women [38–40]. Two features warrant careful analysis. First, harassment is a family of behaviours varying in frequency and severity, from chronic low-level sexist remarks to discrete acts of unwanted sexual attention and coercion; chronic and acute variants may relate to burnout through distinct pathways, with chronic exposure functioning as an ongoing stressor and acute exposure as a threat to mental wellbeing [41,42]. Second, given the strong gender asymmetry in exposure, the harassment-burnout relationship may differ in magnitude and mechanism between men and women. We therefore administered the Sexual Experiences Questionnaire-Department of Defense short form (SEQ-DoD-s) [43] as an optional module, and planned analyses in the subsample to test whether harassment predicted burnout above the primary predictors, whether the four subscales differed in their direct and indirect relationships with burnout, and whether mental wellbeing mediated the harassment-burnout association in women (Hypothesis 5).

## Materials and methods

### Design and participants

Data were collected via an online survey hosted on Qualtrics for eight weeks between May and July 2022. Participants were recruited from the UK film and television workforce, including freelance, contracted, and salaried workers across the industry. A cross-sectional, correlational survey design was used. Eligible participants were aged 18 or over and had worked in the UK film or television industry in any capacity during the preceding 12 months. Recruitment was conducted through the lead author’s professional contacts, social media, and direct approaches to industry guilds, unions, agencies, production companies, and other industry organisations; approximately 1,300 individuals and organisations were contacted directly.

The final sample comprised 415 individuals across more than 100 distinct job titles spanning casting, pre-production, production, post-production, distribution, and performance. Mean age was 38.4 years (SD = 10.8, range 20-79). One hundred eighty-four identified as men (including transgender men), 219 as women (including transgender women), 8 as non-binary, and 4 as self-identifying outside these categories; 8 reported a current gender identity that did not match their sex assigned at birth. The majority were freelance or contracted (n = 339, 81.7%), with 60 (14.5%) salaried and the remainder in mixed arrangements, company owners, or students. London was the most common region (n = 241, 58.1%). Self-reported working hours averaged 52.6 per week (SD = 21.8, median 55); 66.5% exceeded 48 hours, 30.6% exceeded 60. Just under a quarter (24.6%) reported a disability or long-term health condition, and 53.3% had considered leaving the industry in the preceding year. The optional harassment module was completed by 297 participants; for gender-stratified analyses the subsample comprised 115 men and 171 women (N = 286). Non-binary and self-identifying participants were not analysed separately because sample sizes were too small for stable inference. A detailed breakdown of demographics and job titles by department is provided in the S1 Appendix. As the survey was anonymous and open-access, information on individuals who viewed but did not complete the survey, or completed only part of it, cannot be reported. The optional harassment module was completed by 71.6% of participants (297 of 415).

### Materials

The questionnaire was hosted on Qualtrics and comprised an information and consent page, a demographics section, and five validated scales. The harassment scale was presented last, preceded by a second consent screen indicating it was optional and warning of sensitive content.

The 12-item Short Form 12 Health Survey (SF-12) [44] was administered and scored using the algorithm documented in the second edition scoring manual, producing the Physical Component Summary (PCS) and Mental Component Summary (MCS). Each was calibrated to a mean of 50 and SD of 10 in US norms, which are reported to be comparable to UK norms [45]; higher scores indicate better health. The SF-12 has established reliability and validity across multiple populations [44,45].

The 9-item Stigma-9 Questionnaire (STIG-9) [33] measures expectations of negative beliefs and behaviour towards mentally ill people on a 4-point scale (0 = disagree, 1 = somewhat disagree, 2 = somewhat agree, 3 = agree); sums range from 0 to 27, with higher scores indicating greater perceived public stigma (α = .91).

The 11-item European Intrinsic Job Quality Index (EIJQI) [17] measures autonomy (3 items), interaction with colleagues (4 items), intensity (2 items, reverse-coded for the total), and meaningfulness (2 items). Each item was scored from 0 (strongly disagree) to 4 (strongly agree), such that subscale sums range from 0 to four times the number of items, and the EIJQI total ranges from 0 to 44 with higher scores indicating higher intrinsic job quality. Internal consistency in the present sample (α = .81) was higher than that reported by Cascales Mira [17] (α = .72).

The 16-item Maslach Burnout Inventory General Survey (MBI-GS) [9] measures emotional exhaustion (5 items), cynicism (5 items), and professional efficacy (6 items, reverse-coded for the total) on a 7-point frequency scale (0 = never to 6 = every day); higher totals indicate greater burnout (α = .88).

The 16-item Sexual Experiences Questionnaire-Department of Defense short form (SEQ-DoD-s) [43] measures workplace exposure to sexist behaviour, crude or offensive behaviour, unwanted sexual attention, and sexual coercion (4 items each) over 12 months on a 5-point frequency scale (0 = never to 4 = very often), calibrated for greatest precision on moderate-to-severe experiences (α = .95 present sample vs .92 in Stark et al. [43]).

### Procedure and analytic strategy

The study received ethical approval from Northumbria University Ethics Board (Reference 45697, April 2022). All participants provided written informed consent prior to completing the survey. Participants were presented with an information sheet describing the study, the topics covered, the optional harassment section, the four-week withdrawal window, and links to support organisations. After providing informed consent they completed the demographics section, then the SF-12, EIJQI, MBI-GS, and STIG-9, and, optionally, the SEQ-DoD-s. A debrief screen listed support organisations. Anonymised data and analytic syntax are openly deposited on Zenodo [46].

Analyses were conducted in SPSS Version 28 and verified in RStudio using R Version 4.6.1. Primary analyses on the full sample comprised descriptive statistics and assumption checks, bivariate Pearson correlations, and multiple regression of the MBI-GS total and its three subscales on the four primary predictors, with multicollinearity checked via variance inflation factors. Two augmented regressions addressed the secondary aims: (a) a model adding working hours, and (b) a model replacing the EIJQI total with its four subdimensions. Secondary analyses compared women and men, and workers with and without a disability, using Welch’s t-tests with Cohen’s d as the effect size, and Benjamini-Hochberg correction within each comparison family [47]. In the harassment subsample, we ran gender-stratified regressions, a fully adjusted gender X harassment moderated regression with the four primary predictors as covariates, Fisher’s r-to-z tests for gender differences in harassment-burnout and harassment-mental wellbeing correlations [48], and a mediation analysis of the harassment-exhaustion relationship in women with 5,000 bootstrap resamples [49]. Mann-Whitney U tests on the skewed harassment subscales produced the same pattern as parametric tests; only the latter are reported. An a priori G*Power [50] analysis (α = .05, power = .80, f² = .15) required N = 92 for five predictors and N = 103 for seven predictors, comfortably exceeded in both samples.

To mitigate potential sources of bias, the survey was distributed through multiple professional networks and industry bodies rather than a single channel, reducing the risk that participants were drawn from a narrow subset of the workforce. The optional harassment module was administered after the main scales to minimise priming effects on earlier responses. Potential common-method bias from self-report data is acknowledged in the Limitations.

Missing data were handled using listwise deletion. Participants with missing values on any variable included in a given analysis were excluded from that analysis only; the harassment subsample consisted of 297 participants who completed the optional module, with 286 included in gender-stratified analyses after excluding non-binary and self-identifying participants due to small cell sizes.

The study is reported in accordance with the STROBE statement for cross-sectional studies [51].

## Results

### Sample characteristics and working conditions

To contextualise the regression results, we first examined absolute levels of mental wellbeing, working hours, and burnout thresholds. Mean MCS of 36.0 (SD = 11.8) sat below the general-population norm of 50 [44,45], with 82% scoring below 50 and 68% below 42, a threshold commonly used to indicate probable depressive symptoms [52]. Mean PCS was close to the population norm (51.7, SD = 8.9). Applying Schaufeli and Enzmann’s [26] MBI-GS cut-offs, 82% scored in the high-exhaustion range, 72% in the high-cynicism range, and 23% in the low professional-efficacy range. Mean working week was 52.6 hours (median 55), with 66.5% exceeding 48 hours and 30.6% exceeding 60. More than half (53.3%) had considered leaving the industry in the preceding year.

### Descriptive statistics and parametric assumptions

Descriptive statistics are presented in Table 1. For the full sample, all absolute skewness and kurtosis values fell within the ±2 and ±7 thresholds recommended for assuming normality at this sample size [53,54]. The SEQ-DoD-s showed positive skew in the subsample, as is typical for harassment measures. Variance inflation factors across all regression models below were under 1.6, indicating no multicollinearity problems.

**Table 1.**
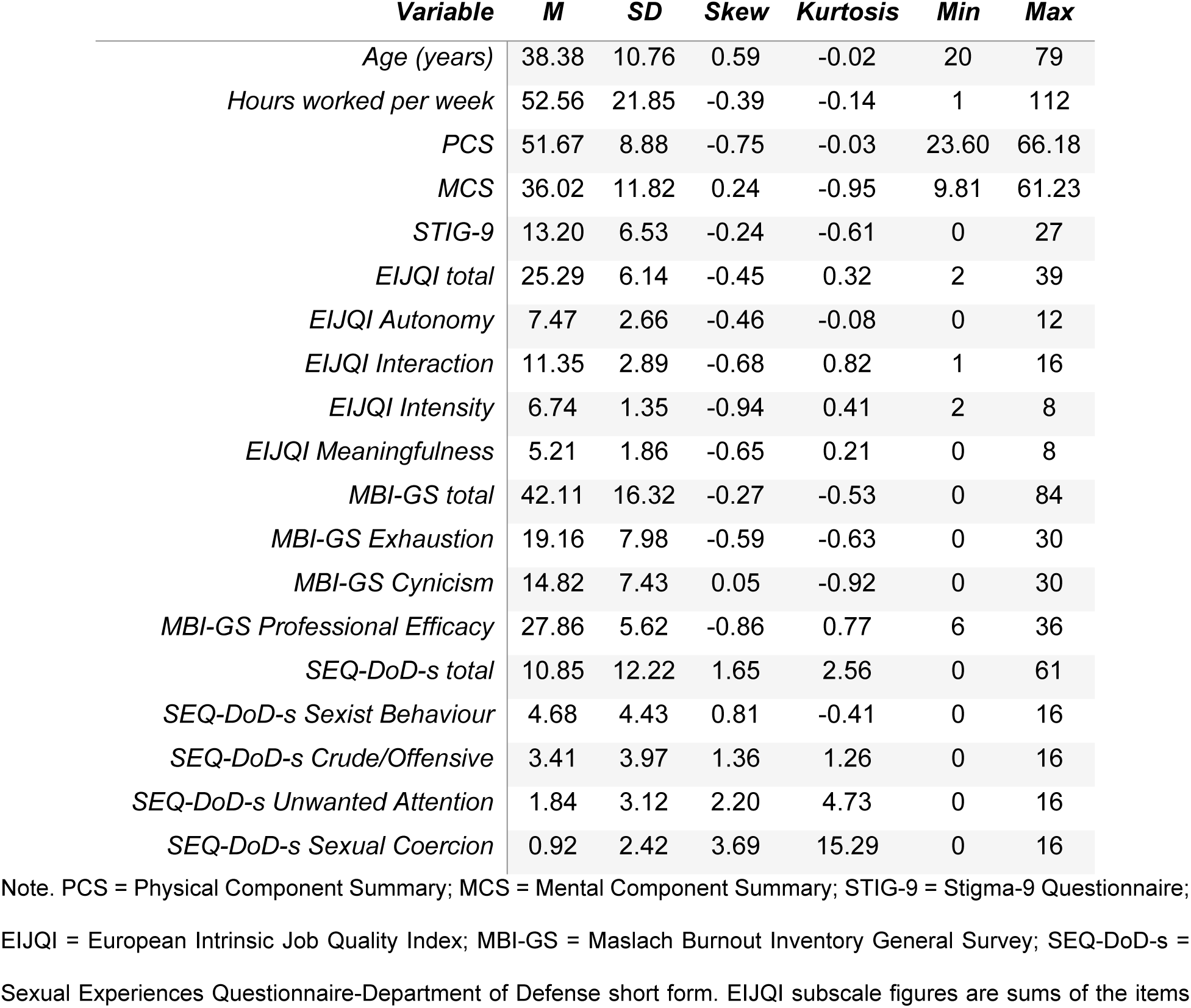
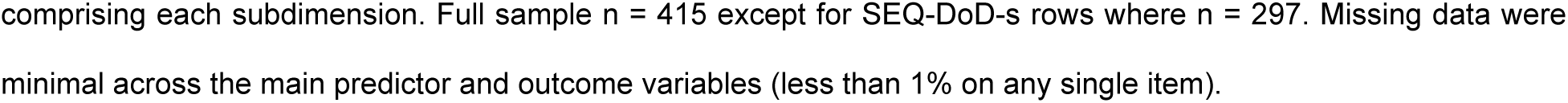
Descriptive statistics for all measured variables.

| *Variable* | *M* | *SD* | *Skew* | *Kurtosis* | *Min* | *Max* |
| --- | --- | --- | --- | --- | --- | --- |
| *Age (years)* | 38.38 | 10.76 | 0.59 | -0.02 | 20 | 79 |
| *Hours worked per week* | 52.56 | 21.85 | -0.39 | -0.14 | 1 | 112 |
| *PCS* | 51.67 | 8.88 | -0.75 | -0.03 | 23.60 | 66.18 |
| *MCS* | 36.02 | 11.82 | 0.24 | -0.95 | 9.81 | 61.23 |
| *STIG-9* | 13.20 | 6.53 | -0.24 | -0.61 | 0 | 27 |
| *EIJQI total* | 25.29 | 6.14 | -0.45 | 0.32 | 2 | 39 |
| *EIJQI Autonomy* | 7.47 | 2.66 | -0.46 | -0.08 | 0 | 12 |
| *EIJQI Interaction* | 11.35 | 2.89 | -0.68 | 0.82 | 1 | 16 |
| *EIJQI Intensity* | 6.74 | 1.35 | -0.94 | 0.41 | 2 | 8 |
| *EIJQI Meaningfulness* | 5.21 | 1.86 | -0.65 | 0.21 | 0 | 8 |
| *MBI-GS total* | 42.11 | 16.32 | -0.27 | -0.53 | 0 | 84 |
| *MBI-GS Exhaustion* | 19.16 | 7.98 | -0.59 | -0.63 | 0 | 30 |
| *MBI-GS Cynicism* | 14.82 | 7.43 | 0.05 | -0.92 | 0 | 30 |
| *MBI-GS Professional Efficacy* | 27.86 | 5.62 | -0.86 | 0.77 | 6 | 36 |
| *SEQ-DoD-s total* | 10.85 | 12.22 | 1.65 | 2.56 | 0 | 61 |
| *SEQ-DoD-s Sexist Behaviour* | 4.68 | 4.43 | 0.81 | -0.41 | 0 | 16 |
| *SEQ-DoD-s Crude/Offensive* | 3.41 | 3.97 | 1.36 | 1.26 | 0 | 16 |
| *SEQ-DoD-s Unwanted Attention* | 1.84 | 3.12 | 2.20 | 4.73 | 0 | 16 |
| *SEQ-DoD-s Sexual Coercion* | 0.92 | 2.42 | 3.69 | 15.29 | 0 | 16 |

Note. PCS = Physical Component Summary; MCS = Mental Component Summary; STIG-9 = Stigma-9 Questionnaire; EIJQI = European Intrinsic Job Quality Index; MBI-GS = Maslach Burnout Inventory General Survey; SEQ-DoD-s = Sexual Experiences Questionnaire-Department of Defense short form. EIJQI subscale figures are sums of the items
comprising each subdimension. Full sample n = 415 except for SEQ-DoD-s rows where n = 297. Missing data were minimal across the main predictor and outcome variables (less than 1% on any single item).

### Bivariate correlations

Pearson correlations are shown in Table 2. Mental wellbeing and intrinsic job quality showed strong negative correlations with overall burnout, while perceived public stigma correlated positively at moderate strength. Physical health correlated weakly with overall burnout but more with emotional exhaustion specifically. Working hours correlated positively with burnout, with the relationship strongest for emotional exhaustion. In the harassment subsample, the SEQ-DoD-s total correlated positively with burnout and negatively with both mental wellbeing and intrinsic job quality.

**Table 2.** Pearson correlations among study variables.

| *Variable* | *1* | *2* | *3* | *4* | *5* | *6* | *7* |
| --- | --- | --- | --- | --- | --- | --- | --- |
| *1. PCS* | - |  |  |  |  |  |  |
| *2. MCS* | -.10* | - |  |  |  |  |  |
| *3. STIG-9* | -.04 | -.30** | - |  |  |  |  |
| *4. EIJI total* | .08 | .44** | -.22** | - |  |  |  |
| *5. Hours per week* | -.11* | -.10* | .02 | -.23** | - |  |  |
| *6. SEQ-DoD-s* | -.15* | -.21** | .23** | -.22** | .08 | - |  |
| *7. MBI-GS total* | -.09 | -.62** | .30** | -.63** | .21** | .22** | - |

Note. Full sample n = 415; n = 297 for correlations involving SEQ-DoD-s. \* p < .05. \*\* p < .01.

### Primary regression analyses

Multiple regression of the MBI-GS total on the four primary predictors yielded a significant model accounting for 56% of the variance, R² = .56, F(4, 410) = 132.02, p < .001. As shown in Table 3, mental wellbeing and intrinsic job quality were the dominant predictors, with comparable standardised coefficients. Physical health made a smaller but significant contribution, and perceived public stigma a small marginally significant one. Parallel regressions on the three MBI-GS subscales showed predictor effects were not uniformly distributed: physical health contributed most to emotional exhaustion, while professional efficacy was predicted primarily by intrinsic job quality and mental wellbeing.

**Table 3.** Multiple regression coefficients predicting MBI-GS total and subscales.

| Predictor | MBI-GS Total |  |  | Exhaustion |  |  | Cynicism |  |  | Prof. Efficacy |  |  |
| --- | --- | --- | --- | --- | --- | --- | --- | --- | --- | --- | --- | --- |
| | $\beta$ | 95% CI | p | $\beta$ | 95% CI | p | $\beta$ | 95% CI | p | $\beta$ | 95% CI | p |
| PCS | -.10 | [-.16, -.03] | .003 | -.17 | [-.25, -.09] | <.001 | -.06 | [-.14, .01] | .107 | -.04 | [-.12, .05] | .394 |
| MCS | -.43 | [-.50, -.35] | <.001 | -.41 | [-.50, -.32] | <.001 | -.36 | [-.45, -.27] | <.001 | .18 | [.09, .28] | <.001 |
| STIG-9 | .07 | [.00, .14] | .047 | .06 | [-.02, .14] | .113 | .08 | [.00, .16] | .060 | -.01 | [-.09, .08] | .899 |
| EIJQI | -.42 | [-.50, -.35] | <.001 | -.28 | [-.36, -.19] | <.001 | -.34 | [-.43, -.25] | <.001 | .39 | [.29, .48] | <.001 |
| $R^2$ | .56 | | | .39 | | | .39 | | | .25 | | |
| $F(4, 410)$ | 132.02 | | | 66.87 | | | 64.83 | | | 33.58 | | |

Note. $N = 415$ . Standardised regression coefficients shown with 95% confidence intervals. All four-predictor models significant at $p < .001$ .

### Working hours and job quality subdimensions

Two augmented regressions extended the primary analysis. First, adding working hours per week to the four-predictor model (Table 4) produced minimal change at the level of the MBI-GS total but a substantial and highly significant effect on emotional exhaustion specifically, making hours the second-largest predictor of exhaustion after mental wellbeing. The association was robust to controlling for physical health, mental wellbeing, perceived stigma, and intrinsic job quality. Hours did not predict cynicism but showed an unexpected positive coefficient on professional efficacy.

**Table 4.** Multiple regression coefficients with working hours added to the primary predictors.

| *Predictor* | *MBI-GS Total* |  |  | *Exhaustion* |  |  | *Cynicism* |  |  | *Prof. Efficacy* |  |  |
| --- | --- | --- | --- | --- | --- | --- | --- | --- | --- | --- | --- | --- |
| | $\beta$ | 95% CI | p | $\beta$ | 95% CI | p | $\beta$ | 95% CI | p | $\beta$ | 95% CI | p |
| *PCS* | -.09 | [-.16, -.03] | .006 | -.15 | [-.22, -.07] | <.001 | -.06 | [-.14, .02] | .129 | -.02 | [-.10, .07] | .650 |
| *MCS* | -.42 | [-.50, -.35] | <.001 | -.40 | [-.49, -.32] | <.001 | -.36 | [-.44, -.27] | <.001 | .19 | [.09, .29] | <.001 |
| *STIG-9* | .07 | [.00, .14] | .039 | .07 | [.00, .15] | .062 | .08 | [-.00, .16] | .056 | .00 | [-.09, .09] | .977 |
| *EIJQI Total* | -.41 | [-.48, -.34] | <.001 | -.23 | [-.32, -.15] | <.001 | -.33 | [-.42, -.25] | <.001 | .42 | [.33, .52] | <.001 |
| *Hours/week* | .06 | [.00, .13] | .065 | .22 | [.14, .29] | <.001 | .03 | [-.05, .11] | .403 | .17 | [.09, .26] | <.001 |
| *R<sup>2</sup>* | .57 |  |  | .44 |  |  | .39 |  |  | .27 |  |  |
| *F(5, 409)* | 106.92 |  |  | 64.05 |  |  | 51.97 |  |  | 30.99 |  |  |

Note. $n = 415$ . Standardised regression coefficients shown with 95% confidence intervals. All five-predictor models significant at $p < .001$ . Hours per week shown as a sixth column relative to Table 3; the four primary predictor coefficients are minimally changed by its inclusion.

Second, replacing the EIJQI total with its four subdimensions identified meaningfulness of the work as the dominant component of the job-quality effect (Table 5). Meaningfulness had the largest standardised coefficient, followed by interaction with colleagues. Autonomy lost significance once meaningfulness was in the model. Intensity carried a significant positive coefficient. The disaggregated seven-predictor model accounted for 61% of the variance in burnout, an increase of 5 percentage points over the four-predictor model. The R² change from Model 1 to Model 3 was statistically significant and tested using the F-test for nested models, F(3, 407) = 18.01, p < .001, and both AIC and BIC strongly preferred [55] the disaggregated model (ΔAIC = 45.7; ΔBIC = 33.6). The four EIJQI subdimensions therefore capture variance in burnout beyond what the composite EIJQI score captures.

**Table 5.** Disaggregated regression with EIJQI subdimensions predicting MBI-GS total and subscales.

| *Predictor* | *MBI-GS Total* |  |  | *Exhaustion* |  |  | *Cynicism* |  |  | *Prof. Efficacy* |  |  |
| --- | --- | --- | --- | --- | --- | --- | --- | --- | --- | --- | --- | --- |
| | $\beta$ | 95% CI | p | $\beta$ | 95% CI | p | $\beta$ | 95% CI | p | $\beta$ | 95% CI | p |
| *PCS* | -.10 | [-.16, -.03] | .003 | -.16 | [-.24, -.08] | <.001 | -.06 | [-.14, .01] | .110 | -.03 | [-.11, .05] | .420 |
| *MCS* | -.40 | [-.47, -.33] | <.001 | -.40 | [-.49, -.32] | <.001 | -.33 | [-.42, -.25] | <.001 | .15 | [.06, .24] | <.001 |
| *STIG-9* | .07 | [.01, .14] | .024 | .06 | [-.02, .14] | .141 | .08 | [.01, .16] | .037 | -.02 | [-.10, .06] | .585 |
| *EIJQI Autonomy* | -.06 | [-.14, .01] | .088 | -.06 | [-.15, .04] | .231 | -.04 | [-.13, .05] | .364 | .05 | [-.04, .15] | .250 |
| *EIJQI Interaction* | -.12 | [-.20, -.04] | .002 | -.11 | [-.21, -.02] | .016 | -.08 | [-.17, .01] | .086 | .08 | [-.01, .18] | .091 |
| *EIJQI Intensity* | .11 | [.05, .17] | <.001 | .24 | [.16, .32] | <.001 | .07 | [-.01, .14] | .070 | .12 | [.04, .20] | .002 |
| *EIJQI Meaningfulness* | -.38 | [-.46, -.31] | <.001 | -.14 | [-.22, -.05] | .002 | -.35 | [-.43, -.26] | <.001 | .46 | [.37, .55] | <.001 |
| *R<sup>2</sup>* | .61 |  |  | .43 |  |  | .43 |  |  | .40 |  |  |
| *F(7, 407)* | 92.54 |  |  | 43.57 |  |  | 44.72 |  |  | 38.00 |  |  |

Note. $n = 415$ . Standardised regression coefficients shown for a model in which the EIJQI total has been replaced by its four subdimensions. Working hours were included in supplementary models reported in the text; for parsimony of presentation, this table reports the seven-predictor model with EIJQI subdimensions but without hours. The hours-only addition produces a comparable pattern with similar coefficients on the other predictors.

### Gender differences

Welch’s t-tests comparing women and men are reported in Table 6, with p-values adjusted for multiple testing using the Benjamini-Hochberg procedure within the comparison family. Women scored lower than men on mental wellbeing, lower on intrinsic job quality (with the largest subscale difference on autonomy), and higher on overall burnout, with the burnout difference concentrated in emotional exhaustion. Small marginal differences appeared on physical health and perceived stigma but did not survive correction. In the harassment subsample (n = 115 men, n = 171 women), women reported greater exposure on every SEQ-DoD-s subscale, with the largest gender difference on sexist behaviour. Women were descriptively more likely than men to have considered leaving the industry (58.4% vs 48.9%), χ²(1, N = 403) = 3.66, p = .056, φ = .10.

**Table 6.** Gender differences on study variables.

| Variable | Men |  | Women |  | t | p | d [95% CI] |
| --- | --- | --- | --- | --- | --- | --- | --- |
|  | M | SD | M | SD |  |  |  |
| Age | 41.10 | 10.29 | 36.64 | 10.70 | 4.25 | <.001<sup>‡</sup> | 0.43 [0.23, 0.62] |
| Hours per week | 52.83 | 20.08 | 52.05 | 23.57 | 0.36 | .721<sup>†</sup> | 0.04 [-0.16, 0.23] |
| PCS | 52.54 | 8.30 | 50.80 | 9.30 | 1.99 | .047<sup>†</sup> | 0.20 [0.00, 0.39] |
| MCS | 38.77 | 11.41 | 34.01 | 11.86 | 4.10 | <.001<sup>‡</sup> | 0.41 [0.21, 0.61] |
| STIG-9 | 12.40 | 6.76 | 13.75 | 6.17 | -2.08 | .038<sup>†</sup> | -0.21 [-0.41, -0.01] |
| EIJQI | 26.11 | 4.94 | 24.67 | 6.90 | 2.43 | .028<sup>‡</sup> | 0.24 [0.04, 0.43] |
| MBI-GS Total | 39.47 | 15.53 | 44.05 | 16.81 | -2.84 | .011<sup>‡</sup> | -0.28 [-0.48, -0.09] |
| MBI-GS Exhaustion | 17.61 | 7.68 | 20.24 | 8.09 | -3.34 | .002<sup>‡</sup> | -0.33 [-0.53, -0.14] |
| *MBI-GS Cynicism* | 14.13 | 7.22 | 15.28 | 7.54 | -1.56 | .120<sup>†</sup> | -0.16 [-0.35, 0.04] |
| *MBI-GS Prof. Efficacy* | 28.27 | 5.38 | 27.47 | 5.89 | 1.43 | .152<sup>†</sup> | 0.14 [-0.05, 0.34] |
| *SEQ-DoD-s Total* | 5.77 | 7.85 | 13.90 | 13.35 | -6.47 | <.001<sup>‡</sup> | -0.71 [-0.95, -0.47] |
| *SEQ-DoD-s Sexist Beh.* | 2.04 | 2.70 | 6.24 | 4.48 | -9.88 | <.001<sup>‡</sup> | -1.09 [-1.34, -0.84] |
| *SEQ-DoD-s Crude/Off.* | 2.22 | 2.66 | 4.09 | 4.37 | -4.51 | <.001<sup>‡</sup> | -0.50 [-0.74, -0.26] |
| *SEQ-DoD-s Unwanted Att.* | 0.97 | 2.17 | 2.37 | 3.50 | -4.18 | <.001<sup>‡</sup> | -0.46 [-0.70, -0.22] |
| *SEQ-DoD-s Coercion* | 0.55 | 1.57 | 1.20 | 2.88 | -2.46 | .028<sup>‡</sup> | -0.27 [-0.50, -0.03] |

Note. Equal variances not assumed, Welch's independent-samples t-tests; full sample $n = 184$ men, 219 women, except SEQ-DoD-s rows where $n = 115$ men, 171 women. Negative $d$ indicates women scored higher. $p$ -values were adjusted for multiple testing using the Benjamini-Hochberg procedure separately within the main comparison family ( $k = 10$ ) and the SEQ-DoD-s family ( $k = 5$ ). <sup>‡</sup> indicates significance surviving BH correction; <sup>†</sup> indicates uncorrected significance only.

Because men in the sample were 4.5 years older on average than women (*M* = 41.1 vs *M* = 36.6), we tested whether the gender differences observed reflected age rather than gender per se. Age adjustment was performed on the five outcomes of which the uncorrected gender difference survived the Benjamini-Hochberg correction (mental wellbeing, overall burnout, emotional exhaustion, EIJQI total, and autonomy); marginal differences on physical health and perceived stigma that had not survived correction were not tested further. Each of the key outcomes was regressed on gender with age entered as a covariate. Gender effects on mental wellbeing (*b* = –3.57, *p* = .002), overall burnout (*b* = +3.26, *p* = .046), emotional exhaustion (*b* = +1.97, *p* = .013), and autonomy (*b* = – 0.74, *p* = .006) were attenuated but remained significant after age adjustment; the gender effect on the EIJQI total became marginal (*b* = –1.20, *p* = .054). Age itself was independently associated with better mental wellbeing (*b* = +0.27 per year, *p* < .001) and lower overall burnout (*b* = –0.30 per year, *p* < .001) across the sample, indicating that older workers in this industry reported lower burnout than younger workers, independent of gender.

### Disability as a risk factor

One hundred and two participants (24.6%) reported a disability or long-term health condition, 300 (72.3%) reported none, and 13 (3.1%) preferred not to say. Comparing the first two groups (Table 7), workers with a disability reported poorer physical health, modestly poorer mental wellbeing, higher overall burnout, and higher emotional exhaustion. The most striking contrast was on perceived public stigma: workers with a disability scored notably higher on the STIG-9, suggesting they anticipated greater public stigma about mental ill-health than non-disabled peers. Intrinsic job quality did not differ significantly between groups.

**Table 7.**
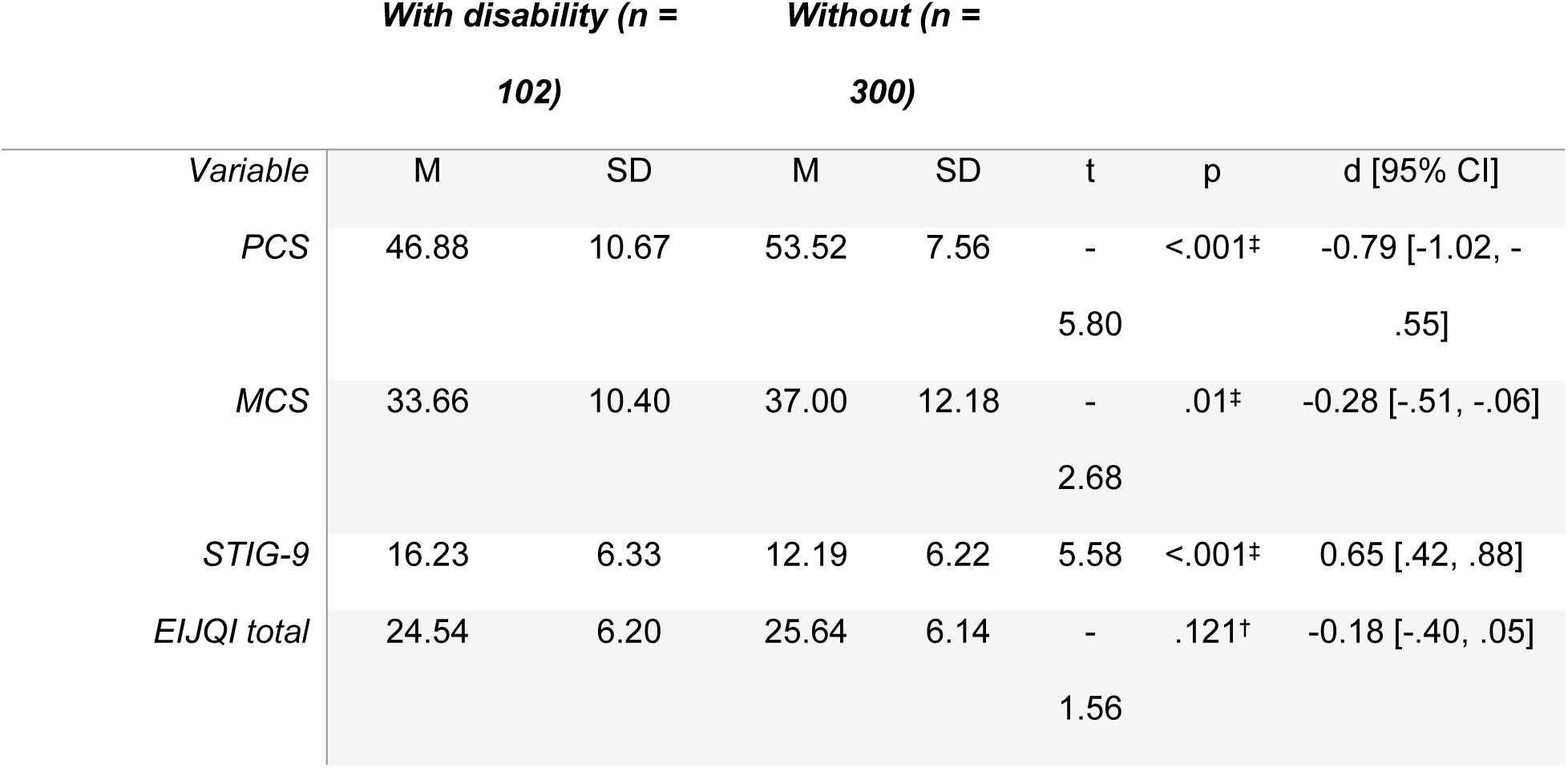
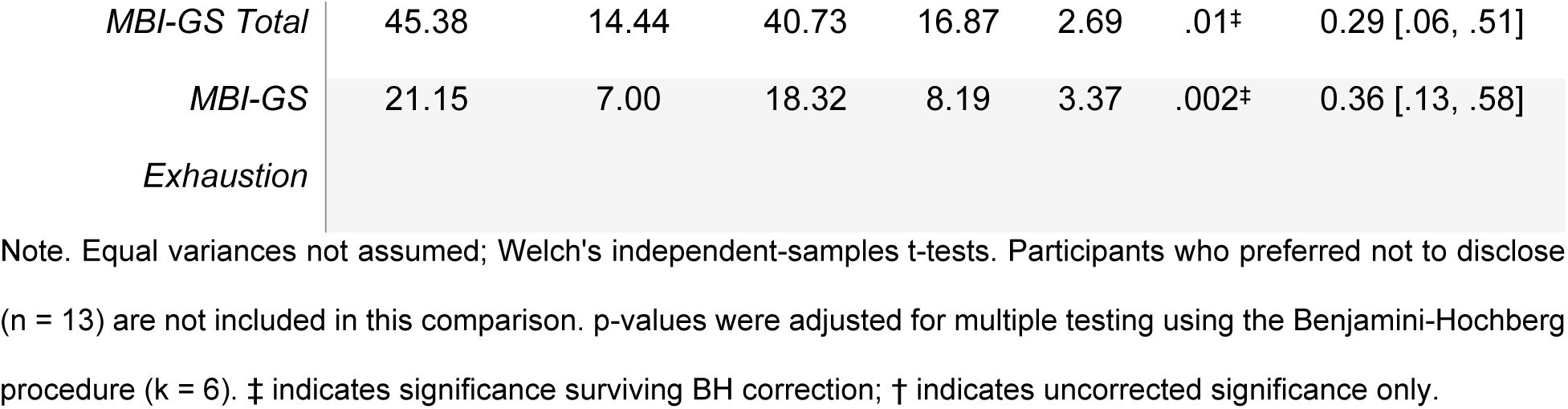
Comparison of workers with and without a self-reported disability or long-term health condition.

### Harassment, burnout, and mediation by mental wellbeing

In gender-stratified regressions with all five predictors, the SEQ-DoD-s total did not add significant predictive variance to burnout in either subsample. A fully adjusted moderated regression, with the four primary predictors as covariates, returned a non-significant SEQ-DoD-s X gender interaction on the MBI-GS total (p = .22); the harassment-burnout association is therefore not significantly stronger in women than in men at the overall burnout level. However, in women considered separately, harassment correlated significantly with both burnout (r = .25, p = .001) and poorer mental wellbeing (r = –.18, p = .019); the corresponding correlations in men were not significant (burnout r = .06, p = .537; mental wellbeing r = –.14, p = .151). Fisher’s r-to-z tests indicated that the harassment-exhaustion correlation was significantly stronger in women than in men (z = 2.19, p = .029), while gender differences on the harassment-burnout total and harassment-mental wellbeing correlations were not statistically significant (z = 1.58, p = .115 and z = 0.37, p = .711 respectively). A mediation analysis in women showed that mental wellbeing mediated the harassment-exhaustion effect: total effect b = 0.16, p < .001; direct effect b = 0.11, p = .003; indirect effect ab = 0.05, bootstrapped 95% CI [0.01, 0.09], accounting for approximately 30% of the total effect. The same pattern obtained for the MBI-GS total: total b = 0.29, p = .001; direct b = 0.17, p = .022; indirect ab = 0.12, 95% CI [0.03, 0.23], accounting for approximately 42% of the total effect. Mediation was not tested in men, since neither the total effect nor the harassment-to-mental-wellbeing path was significant.

Subscale-level analyses in women (Table 8) showed that of the four SEQ-DoD-s components, only sexist behaviour retained a significant association with emotional exhaustion after controlling for all four primary predictors. Crude or offensive behaviour showed a non-significant trend; unwanted sexual attention and sexual coercion, present in bivariate analyses, were not significant in the fully controlled model. In women, chronic low-level sexist behaviour was therefore associated with exhaustion both directly and indirectly through impaired mental wellbeing, while more severe but less frequent forms showed evidence of indirect effects only.

**Table 8.** Sexual harassment subscales as predictors of emotional exhaustion in women, controlling for primary predictors.

| *SEQ-DoD-s Subscale* | $\beta$ | 95% CI | $p$ |
| --- | --- | --- | --- |
| *Sexist Behaviour* | .15 | [.02, .28] | .022 |
| *Crude or Offensive Behaviour* | .12 | [-.01, .25] | .074 |
| *Unwanted Sexual Attention* | .09 | [-.04, .21] | .174 |
| *Sexual Coercion* | .06 | [-.06, .19] | .322 |

Note. $n = 171$ . Each row reports the standardised coefficient for the SEQ-DoD-s subscale after controlling for PCS, MCS, STIG-9, and EIJI in the female subsample. Higher $\beta$ indicates greater exposure on that subscale predicting higher emotional exhaustion.

### Burnout and intent to leave the industry

Participants who had considered leaving in the preceding 12 months (n = 221) scored higher on the MBI-GS than those who had not (n = 194), M = 49.86 vs M = 33.29, t(400.23) = 11.92, p < .001, d = 1.18. In a multivariate logistic regression with all four primary predictors and the MBI-GS total entered simultaneously, burnout remained a significant independent predictor of intent to leave, OR = 2.73 per SD, 95% CI [1.92, 3.94], p < .001, almost tripling the odds. Intrinsic job quality also remained an independent predictor, OR = 0.72 per SD, 95% CI [0.53, 0.97], p = .032. The unique effects of physical health, mental wellbeing, and stigma were not significant once burnout was in the model, consistent with burnout being a proximal mechanism through which upstream stressors translate into workforce withdrawal.

## Discussion

This is, to our knowledge, the first peer-reviewed empirical study of occupational burnout in the UK film and television workforce. Lower mental wellbeing and intrinsic job quality were the dominant predictors, with smaller contributions from physical health and stigma; the four accounted for 56% of the variance, rising significantly to 61% when the EIJQI was disaggregated. Working hours independently predicted exhaustion. Workers with a disability reported higher stigma and exhaustion. In the harassment subsample, the harassment-burnout relationship was present in women but not men, approximately 30% mediated by mental wellbeing, with sexist behaviour directly predicting exhaustion after all controls.

### Mental wellbeing and job quality

The mental wellbeing scores are notable not only for their predictive role but for their absolute level. A mean MCS sitting more than 1.4 standard deviations below the UK general-population norm [45], with 68% scoring below the SF-12 threshold conventionally used to indicate probable depressive symptoms [52], provides validated psychometric confirmation of what the Looking Glass surveys reported through self-rated single items [2,3]. Adjacent UK creative-sector studies have documented elevated depression during the COVID-19 lockdown [7] and qualitative distress across the cultural sector including film and television [8]; the present findings extend this picture to ordinary industry working conditions, suggesting the mental health concerns are structural rather than pandemic-driven.

The strength of the MCS-burnout association is consistent with the broader literature on the close coupling of mental health and burnout [25,28], and the fact that mental wellbeing remained the largest predictor of burnout after controlling for job quality, physical health, and stigma indicates that workforce mental health cannot be reduced to job design alone. The standardised coefficient for intrinsic job quality was nearly identical to that for mental wellbeing, and the disaggregated model (Table 5) located most of this effect in a single subdimension: meaningfulness of the work, the second-largest coefficient in the full model after mental wellbeing itself. Interaction with colleagues added an independent effect, consistent with the protective role of social resources in the JD-R framework [15]. Autonomy lost independent association with burnout once meaningfulness was controlled, suggesting the autonomy-burnout link in this workforce may be largely mediated by the meaning autonomy affords. Intensity carried a significant positive coefficient, indicating higher intensity drives higher burnout independently of other resources.

### Working hours

The specificity of the hours effect, substantial on emotional exhaustion but absent on cynicism, is consistent with hours acting on burnout primarily through fatigue accumulation rather than eroded resources or impaired affect. The descriptive picture and the regression evidence together make a case for working hours as a primary target of industry-level reform, a point reinforced by the retention findings reported below.

### Physical health and perceived stigma

Mean physical health was close to general-population norms, yet the physical health-burnout link was supported, particularly for emotional exhaustion, consistent with bidirectional pathways between physical condition and emotional fatigue [23]. The absence of a larger physical-health decrement may reflect selection: workers no longer physically able to sustain industry hours may leave altogether [6].

Perceived public stigma was a small but significant predictor, and its moderate bivariate correlation with burnout suggests a more substantial association largely captured by mental wellbeing once it is in the model. Stigma appears to operate primarily as an upstream risk factor for poor mental wellbeing rather than exerting a direct effect on burnout. The STIG-9 also measures perceived public stigma rather than internalised self-stigma; instruments such as the Internalised Stigma of Mental Illness scale [56] might capture a more directly burnout-relevant dimension. The disability finding adds an important qualification: workers with a disability or long-term health condition reported elevated stigma, a medium-to-large effect, making this subgroup the most exposed to the stigma pathway and the most likely to benefit from industry initiatives that normalise access to mental health support.

### Sexual harassment, mental wellbeing, and burnout

As anticipated in the analytic plan, the harassment findings distinguish direct from mediated effects. Although the SEQ-DoD-s total did not add unique variance in the fully adjusted regression, this null result is incomplete: in women, approximately 30% of the harassment-exhaustion association was mediated by mental wellbeing. The mechanism is straightforward: harassment damages mental health, and impaired mental health drives exhaustion. Once mental wellbeing enters the regression, variance shared with harassment is partitioned to it, leaving harassment with little residual explanatory power. The harassment effect is real but mostly indirect.

The subscale analyses sharpen the picture: only sexist behaviour, capturing chronic low-level sexist remarks, retained a direct effect on exhaustion in women after all primary predictors were controlled. The other subscales measure rarer but more acute experiences, and their effects on burnout appear to operate fully through mental wellbeing. This is consistent with literature distinguishing chronic everyday sexism from discrete-event severe harassment [41,42]; for an exhaustion-based syndrome in which chronicity is definitional, ongoing low-grade sexism may have a direct fatiguing effect. The absence of any harassment-burnout link in men is consistent with the broader literature [39], and the elevated exhaustion difference in women (compared with the meta-analytic norm reported by Purvanova and Muros [18]) is consistent with industry-specific stressors amplifying this pathway.

### Industry retention and the structural nature of the problem

A median working week of 55 hours is incompatible with the recovery cycles that protect against chronic exhaustion [57]. That 82% of the sample scored in the high-exhaustion range and 72% in the high-cynicism range on the MBI-GS, against population baselines well below 50% in non-helping professions [26], reinforces the point. More than half (53.3%) had considered leaving in the preceding year, and a standard-deviation increase in burnout almost tripled the odds of doing so, consistent with the Film and TV Charity’s retention study [6] and identifying burnout as the proximal mechanism through which working conditions translate into workforce withdrawal.

### Implications, limitations, and future directions

The findings extend JD-R and COR frameworks to a workforce whose project-based structure differs from the salaried samples on which most burnout research has been conducted. The dominant role of meaningfulness is consistent with JD-R’s emphasis on resources as buffers against chronic demands; the comparably strong contribution of mental wellbeing suggests that in a workforce moving across many production environments the cumulative affective toll may be more burnout-relevant than any single project’s features.

Four practical implications follow. Impaired mental wellbeing at this scale requires an occupational-health rather than individual-help-seeking approach, particularly for workers with a disability. Working hours and intensity findings indicate that hour-reduction and recovery-time protection are high-leverage structural reforms. Meaningfulness is the strongest protective resource: preserving creative agency and conditions of perceived meaning is consequential. Harassment interventions should address chronic low-level sexist behaviour alongside more severe forms, since our data show it most directly fatigues women workers. The Film and TV Charity’s Principles for Mentally Healthy Productions, launched in February 2026 following a twelve-month cross-sector consultation [58], converge with our findings. The Principles’ three pillars (People Support, Content Impact, and Work Planning) map respectively onto our mental wellbeing and stigma findings, the meaningfulness subdimension of intrinsic job quality, and the working hours and intensity findings. The convergence of independent industry consultation and empirical psychometric findings on the same structural targets reinforces the case that these are the highest-leverage points for intervention.

Strengths include a large occupationally diverse sample, validated instruments with strong internal consistencies, and openly deposited data. Limitations temper interpretation, the cross-sectional design precludes causal inference; the associations are equally consistent with reverse causation and shared third causes. Non-probability recruitment, unavoidable in a workforce with no sampling frame, may over-represent workers most engaged with industry networks, and the harassment subsample was self-selected. Longitudinal designs would be needed to test the mediational pathways identified, and intervention research with industry bodies would be most informative; an industry-specific psychometric instrument reflecting project-based freelance work would help future studies move beyond instruments designed for salaried samples.

The gender comparisons were adjusted for age given a mean age difference of 4.5 years between men and women in the sample, but residual confounds with career stage, project role distribution, and workforce entry cohort cannot be ruled out.

## Conclusion

In 415 UK film and television professionals, occupational burnout was strongly predicted by mental wellbeing and intrinsic job quality, with meaningfulness of the work as the largest job-quality subdimension; working hours independently predicted emotional exhaustion. Two-thirds scored in the range indicative of probable depression, four in five in the high-exhaustion range, and over half had considered leaving in the past year. Workers with a disability reported elevated stigma; and in women, sexual harassment exposure, particularly chronic low-level sexist behaviour, was associated with burnout both directly and through impaired mental wellbeing. The findings are consistent with an industry-wide occupational health concern requiring structural reform of working hours, harassment culture, and meaningfulness-preserving production practices.

## Data Availability

All data and analytic syntax underlying the findings of this study are publicly available in the Zenodo repository at https://doi.org/10.5281/zenodo.22679502. The deposit includes the anonymised dataset (.RData and CSV formats), a variable codebook, the SPSS syntax used to compute SF-12 MCS and PCS scores, the R analytic script reproducing all reported analyses, and a README documenting file contents and reproduction steps.

## Acknowledgements

The authors wish to thank the UK film and television industry professionals who gave their time to participate in this study.

## Supporting information

**S1 Appendix. Detailed demographics and job titles by department.** Full breakdown of participant demographic characteristics and the distribution of job titles across departments in the UK film and television workforce.

